# Pathways from Social Support to Cognition Among Adults in the United States: A Longitudinal Mediation Analysis of Perceived Stress and Sleep Quality

**DOI:** 10.64898/2026.08.10.26360099

**Authors:** Soomin Ryu, Eunjin L. Tracy, Mary Beth Miller, Jill A. Kanaley, Idethia Shevon Harvey

**Author notes:** Corresponding Author: Soomin Ryu 710 Lewis Hall, Department of Public Health College of Health Sciences, University of Missouri, Columbia, MO, USA, 65211.

## Abstract

**Objective:** We examined whether perceived stress and sleep quality mediate the association between social support and later cognition among adults in the United States.

**Methods:** We used longitudinal Midlife in the United States (MIDUS) data. Social support (1995–1996) was modeled as a latent construct indicated by family and friend support. Perceived stress and sleep quality were measured in the MIDUS 2 Biomarker Project (2004–2009), and cognition was assessed in MIDUS 2 and MIDUS 3. Structural equation models evaluated parallel indirect pathways, adjusting for MIDUS 2 cognition and covariates.

**Results:** Higher social support was associated with lower perceived stress (β=−0.32, 95% CI:−0.41, −0.23) and better sleep quality (β=−0.25, 95% CI:−0.35, −0.15). Greater perceived stress was associated with lower cognition (β=−0.06, 95% CI:−0.11, −0.01), whereas sleep quality was not associated with cognition. Direct and total social support-cognition associations were not statistically significant. A small positive indirect association through perceived stress was identified (β=0.02, 95% CI:0.00, 0.04); no indirect association through sleep quality was identified.

**Conclusions:** Findings are consistent with a possible psychosocial pathway through perceived stress, although the effect was modest and total and direct associations were not statistically significant. Sleep quality showed no statistically significant indirect association.

## 1. INTRODUCTION

Given the rapidly aging U.S. population, cognitive decline represents a major public health challenge.^1^ Alzheimer’s disease and related dementias are among the leading causes of death among older adults^2^ and impose substantial societal and economic burdens.^3^ As cognitive decline predicts dementia progression,^4^ identifying modifiable determinants of cognition is important.

Social relationships are fundamental determinants of health across the lifespan,^5,6^ and social support in particular has emerged as an important protective factor for cognition.^7,8^ Several observational studies have shown that higher social support is associated with better cognition and reduced risk of incident dementia.^8,9^ Supportive relationships may promote cognitive resilience through healthier behaviors, adaptive coping, and reduced neuroendocrine and inflammatory dysregulation.^10,11^ However, the mechanisms linking social support with cognitive trajectories remain insufficiently understood.

The Stress Process Model provides a framework for understanding how social conditions and resources shape health through differential exposure to and appraisal of stressors.^12^ Complementing this perspective, the stress-buffering hypothesis posits that supportive relationships attenuate psychological and physiological responses to stress, thereby reducing their adverse health consequences.^10^ Chronic stress contributes to cumulative neuroendocrine and inflammatory dysregulation associated with allostatic load, which may accelerate biological aging.^13,14^ Together, these frameworks suggest that social support may influence cognitive health through stress-related pathways.

Perceived stress represents one such pathway. Higher stress has been associated with poorer memory, attention, and executive functioning, as well as dysregulation of stress-related neural systems implicated in cognitive aging.^11,15^ Supportive relationships may reduce perceived stress and protect cognition over time.

Sleep quality may represent another pathway linking social support and cognition. Sleep plays a critical role in neural restoration, memory consolidation, and cognitive regulation.^16^ Poor sleep quality has been associated with worse cognitive performance.^17,18^ Social support may promote better sleep through psychosocial mechanisms such as emotional security and reduced loneliness. Perceived stress and sleep quality may therefore represent complementary pathways linking social environments to cognitive aging.^10,11,16^

Although prior studies have examined social resources, sleep, or stress in relation to cognition, few longitudinal studies have simultaneously evaluated stress and sleep quality as parallel mediators of the association between social support and cognition. Prior work examining sleep as a mediator of social resources and cognition has often relied on cross-sectional designs or failed to account for baseline cognition.^19,20^ As a result, the long-term psychosocial pathways linking social support to cognitive trajectories remain unclear.

This study used longitudinal Midlife in the United States (MIDUS) data spanning approximately two decades to test whether perceived stress and sleep quality mediate the association between social support and later cognition among adults in the United States. We examined longitudinal social support and cognitive health and tested psychosocial pathways.

## 2. METHODS

### 2.1. Study design and population

We used data from the MIDUS study, a national longitudinal cohort designed to examine psychosocial, behavioral, and biological determinants of health across adulthood. MIDUS 1 (1995–1996) enrolled non-institutionalized adults. Participants were re-interviewed during MIDUS 2 (2004–2006) and MIDUS 3 (2013–2014). The present analyses integrate data from multiple MIDUS projects. Measures of social support were drawn from the MIDUS 1 Core Survey (1995–1996). Perceived stress and sleep quality were assessed during the MIDUS 2 Biomarker Project (Project 4, 2004–2009). Cognition was assessed at two time points: earlier-wave cognition was measured in the MIDUS 2 Cognitive Project (2004–2006), and follow-up cognition was measured in the MIDUS 3 Cognitive Project (2013–2017).

Figure 1 shows MIDUS subsample contributions to the analytic dataset. The merged dataset included 7,809 participants. Models used full information maximum likelihood (FIML), retaining partially observed data under a missing-at-random assumption.^21^ Participants with data on at least one modeled variable formed the analytic pool; 194 observations missing all modeled variables were excluded during the estimation, yielding 7,615 participants. We included variables associated with attrition and project participation to support this assumption. Complete-case models included 738 participants with non-missing modeled variables.

**Figure 1.**
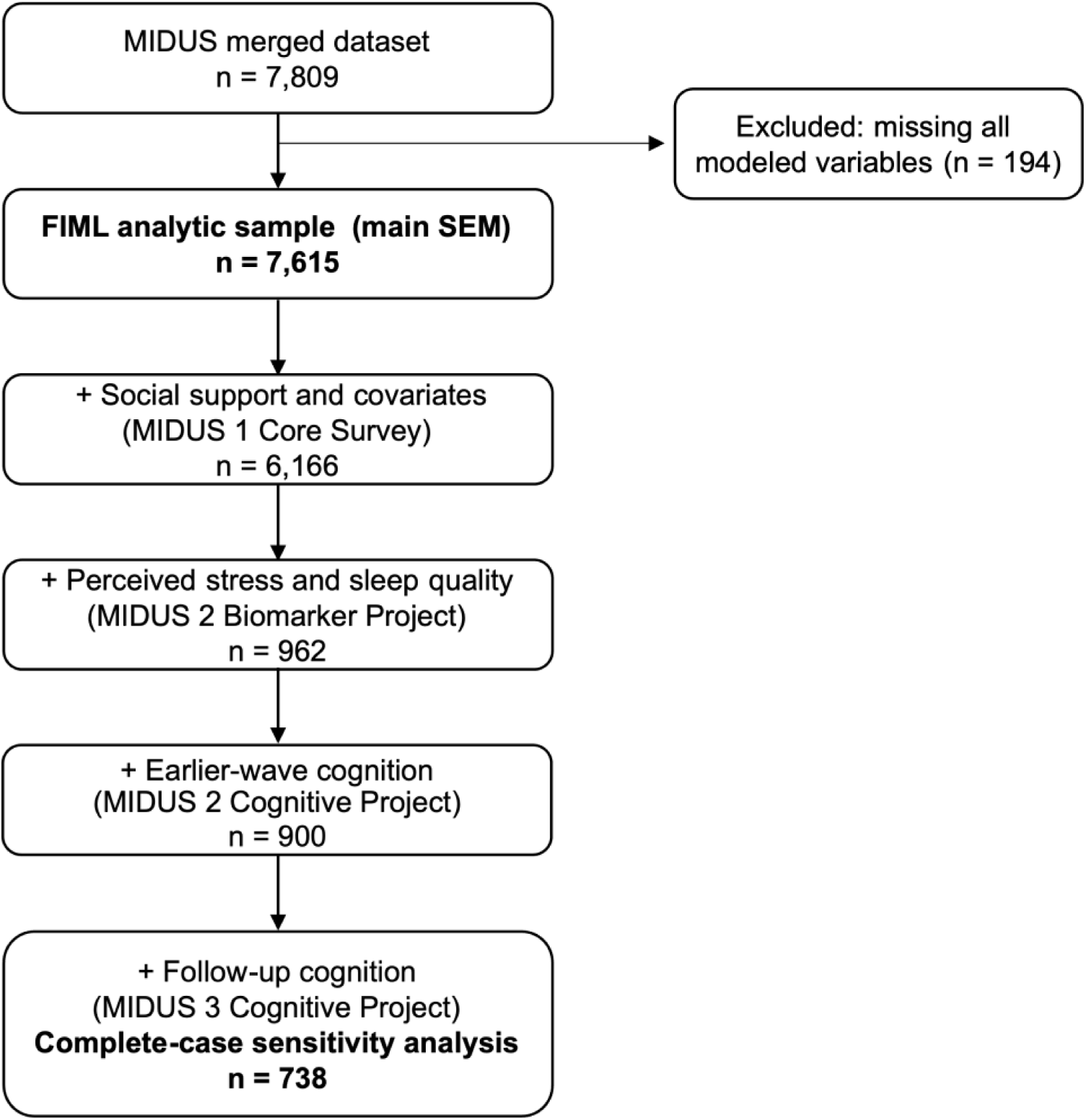
Flow diagram of adults in the United States contributing data across Midlife in the United States study projects, 1995–2017. Notes: Counts below the FIML analytic sample are cumulative complete-data overlaps (the number of participants with non-missing data on all variables required up to that stage), not marginal project-participation counts, and they do not define the estimation sample. The primary SEM was estimated with FIML and included all 7,615 participants with data on at least one endogenous model variable; the complete-case sensitivity analysis included the 738 participants with complete data on all modeled variables. Abbreviation. MIDUS=Midlife in the United States, FIML=full information maximum likelihood, SEM=structural equation modeling

### 2.2. Measures

Social support was assessed using family and friend support scales.^22^ Each domain included four items evaluating emotional support, understanding, reliability, and opportunities for confiding. Response options ranged from 1 (“a lot”) to 4 (“not at all”), and reverse-coded mean scores were calculated so that higher scores indicated greater perceived support (Cronbach’s α=0.82 for family support, α=0.88 for friend support). A latent social support factor was modeled using family and friend support scores as indicators, with higher latent values reflecting greater overall perceived support.

Perceived stress was assessed using the 10-item Perceived Stress Scale (PSS-10).^23^ The PSS evaluates the extent to which individuals perceive their lives as unpredictable, uncontrollable, and overloaded during the past month. Each item was rated on a 5-point frequency scale ranging from 1 (“never”) to 5 (“very often”). Positively worded items were reverse coded so that higher values reflect greater perceived stress. Items were summed to create a composite perceived stress score, with higher scores indicating greater stress (possible range: 10–50). The scale demonstrated good internal consistency in the analytic sample (Cronbach’s α=0.86).

Sleep quality was measured using the Pittsburgh Sleep Quality Index (PSQI), a widely used self-reported instrument that assesses sleep quality and disturbances across seven domains over the past month.^24^ The 19-item PSQI sums the seven components, with higher scores indicating worse sleep quality (possible range: 0–21; Cronbach’s α=0.74) and scores >5 indicating “poor” global sleep quality.

Cognition was assessed using the Brief Test of Adult Cognition by Telephone (BTACT), a well-validated measure designed for large-scale epidemiologic studies.^25^ The BTACT assesses multiple domains of cognitive aging, including episodic memory, executive function, working memory, reasoning, processing speed, and verbal fluency.^26^ Following MIDUS scoring procedures, task scores were combined into a composite score, with higher values indicating better performance. The MIDUS 3 composite score was standardized using the mean and standard deviation of the main national MIDUS 2 sample to facilitate longitudinal comparability across waves. To reduce confounding, we included earlier-wave cognition, measured in the MIDUS 2 Cognitive Project (2004–2006). Among participants with observed perceived stress, sleep quality, and MIDUS 2 and 3 cognition (n=760), MIDUS 2 cognition preceded biomarker collection for all participants; the median interval was 22 months (interquartile range: 11–36; range: 1–60).

Covariates are age, sex, race and ethnicity, partnered status, education, household income, depressive symptoms, and chronic conditions, all measured at MIDUS 1, as well as earlier-wave cognition measured at MIDUS 2. Age and earlier-wave cognition were modeled as continuous variables. Sex, race and ethnicity, partnered status, depressive symptoms, and chronic conditions were represented using binary indicators. Education and household income were modeled as ordered categorical variables, with higher values indicating higher socioeconomic position.

### 2.3. Statistical analysis

We first described the analytic sample using summary statistics for all study variables and examined baseline characteristics according to mediator-data availability. We then specified a structural equation model (SEM) with perceived stress and sleep quality as parallel mediators of the association between social support and follow-up cognition. Social support was modeled as a latent construct indicated by family and friend support. Perceived stress and sleep quality were regressed on latent social support and covariates, and follow-up cognition was regressed on perceived stress, sleep quality, latent social support, earlier-wave cognition, and covariates. Residual errors of perceived stress and sleep quality were allowed to covary because both mediators were assessed concurrently.

The primary SEM was estimated using FIML with robust standard errors. Because perceived stress and sleep quality were assessed in the MIDUS 2 Biomarker Project and cognitive outcomes were assessed in MIDUS cognitive projects, the number of jointly observed participants differed across pathways. Under FIML, all available observations contributed to estimation under a missing-at-random assumption. Because the analyses combined participants from multiple MIDUS projects and waves, sampling weights were not used. All estimates were standardized. We estimated direct, indirect, and total associations using the product-of-coefficients approach. Standard errors and 95% confidence intervals (CI) were calculated using the delta method implemented with nlcom. Standardized root mean squared residual (SRMR) was not reported for the primary robust FIML model because of missing data; it was available for the complete-case model.

We conducted sensitivity analyses to assess the robustness of the findings to missing data, mediator specification, and alternative mediator ordering. First, we re-estimated the primary SEM among participants with complete data on all modeled variables and covariates. Second, we estimated separate complete-case mediator-specific models for perceived stress and sleep quality because the sleep-only FIML model did not converge. Third, we replaced the latent social-support factor with an observed family/friend mean composite and entered family and friend support separately. Fourth, we estimated two sequential mediation models using alternative statistical orderings of perceived stress and sleep quality. Analyses were conducted in Stata/SE 18.5 using sem with method(mlmv) and robust standard errors.

## 3. RESULTS

Table 1 presents the characteristics of the analytic sample. At MIDUS 1, participants had a mean age of 46.4 years, 51.6% were female, and 89.7% identified as non-Hispanic White. Participants perceived relatively strong support from family (mean=3.4) and friends (mean=3.2). At MIDUS 2, perceived stress was moderate (mean=22.2) while sleep quality was poor (mean=6.2), with 48.1% of participants exceeding the PSQI cutoff (>5) for poor sleep quality. Cognition at MIDUS 3 was standardized using the mean and standard deviation of the main national MIDUS 2 sample for longitudinal comparability. Because key variables came from different projects, jointly observed sample sizes varied by path; pairwise overlaps are reported in Supplementary Table 1.

**Table 1.** Characteristics of adults in the United States in the Midlife in the United States study, 1995–2017. a. The cognitive composite scores at MIDUS 3 were standardized using the mean and standard deviation of the MIDUS 2 national sample to facilitate longitudinal comparability across waves. Because MIDUS 3 scores were scaled to the MIDUS 2 reference distribution, the mean MIDUS 3 value reflects the position of the MIDUS 3 sample relative to the MIDUS 2 reference mean rather than a direct estimate of cognitive change over time. Notes: Displayed n values are variable-specific marginal counts, not the overall FIML sample. Ranges shown reflect the observed minimum and maximum values in the analytic sample. Theoretical scale ranges are 10–50 for the perceived stress scale and 0–21 for the sleep quality. Abbreviation. MIDUS=Midlife in the United States, SD=standard deviation, GED=general educational development, FIML=full information maximum likelihood

|  | Mean (SD) | % | n |
| --- | --- | --- | --- |
| Age (observed range: 20 to 75), MIDUS 1 | 46.4 (13.0) |  | 7,049 |
| Sex, MIDUS 1 |  |  | 7,106 |
| Female |  | 51.6 |  |
| Male |  | 48.4 |  |
| Race and ethnicity, MIDUS 1 |  |  | 6,256 |
| Non-Hispanic White |  | 89.7 |  |
| Non-White |  | 10.3 |  |
| Partnered status, MIDUS 1 |  |  | 7,106 |
| Married or cohabiting |  | 70.7 |  |
| Not married or cohabiting |  | 29.3 |  |
| Education, MIDUS 1 |  |  | 7,095 |
| GED / high school graduate or less |  | 38.6 |  |
| Some college |  | 30.6 |  |
| College graduate or higher |  | 30.7 |  |
| Household income, MIDUS 1 |  |  | 6,325 |
| < \$35,000 | | 29.4 | |
| \$35,000–\$74,999 | | 33.9 | |
| \$75,000–\$99,999 | | 10.8 | |
| ≥ \$100,000 | | 26.0 | |
| Depressive symptoms, MIDUS 1 |  |  | 7,108 |
| Yes |  | 13.2 |  |
| No |  | 86.8 |  |
| Chronic conditions, MIDUS 1 |  |  | 6,308 |
| Yes |  | 76.1 |  |
| No |  | 23.9 |  |
| Family support (observed range: 1 to 4, higher=better support), MIDUS 1 | 3.4 (0.6) |  | 6,244 |
| Friend support (observed range: 1 to 4, higher=better support), MIDUS 1 | 3.2 (0.7) |  | 6,244 |
| Perceived stress scale (observed range: 10 to 48, higher=greater stress), MIDUS 2 | 22.2 (6.3) |  | 1,249 |
| Sleep quality (observed range: 0 to 19, higher=poorer sleep quality), MIDUS 2 | 6.2 (3.7) |  | 1,172 |
| Poor sleep quality (score > 5) |  |  | 1,172 |
| Yes |  | 48.1 |  |
| No |  | 51.9 |  |
| Earlier-wave cognition (observed range: -2.9 to 3.6), MIDUS 2 | 0.0 (1.0) |  | 3,973 |
| Cognition (observed range: -2.6 to 2.0), MIDUS 3 <sup>a</sup> | 0.0 (0.7) |  | 3,043 |

Supplementary Table 2 compares baseline characteristics by mediator data availability. Participants with mediator data were more socioeconomically advantaged and had slightly higher cognitive scores than those without mediator data. Partnered status also differed across groups, while age, sex, depressive symptoms, chronic conditions, and social support levels were similar.

Figure 2 summarizes the SEM estimated using FIML, evaluating perceived stress and sleep quality as parallel mediators of the association between social support and subsequent cognition. The latent social support factor loaded significantly on family and friend support. Higher social support was associated with lower perceived stress (β=−0.32, 95% CI: −0.41, −0.23) and lower PSQI scores, indicating better sleep quality (β=−0.25, 95% CI: −0.35, −0.15). In the cognition equation, perceived stress was associated with lower follow-up cognition (β=−0.06, 95% CI: −0.11, −0.01), whereas sleep quality was not statistically associated with cognition. The direct association between social support and follow-up cognition was not statistically significant after accounting for mediators, while baseline cognition was strongly associated with follow-up cognition (β=0.65, 95% CI: 0.62, 0.67). The residual covariance between perceived stress and sleep quality was statistically significant (cov=0.27), indicating shared variance between mediators beyond that explained by social support and covariates. Model fit indices for the primary model were Akaike Information Criterion (AIC)=175,696.5; Bayesian Information Criterion (BIC)=176,431.9; Coefficient of Determination (CD)=0.90; and R² for follow-up cognition=0.71.

**Figure 2.**
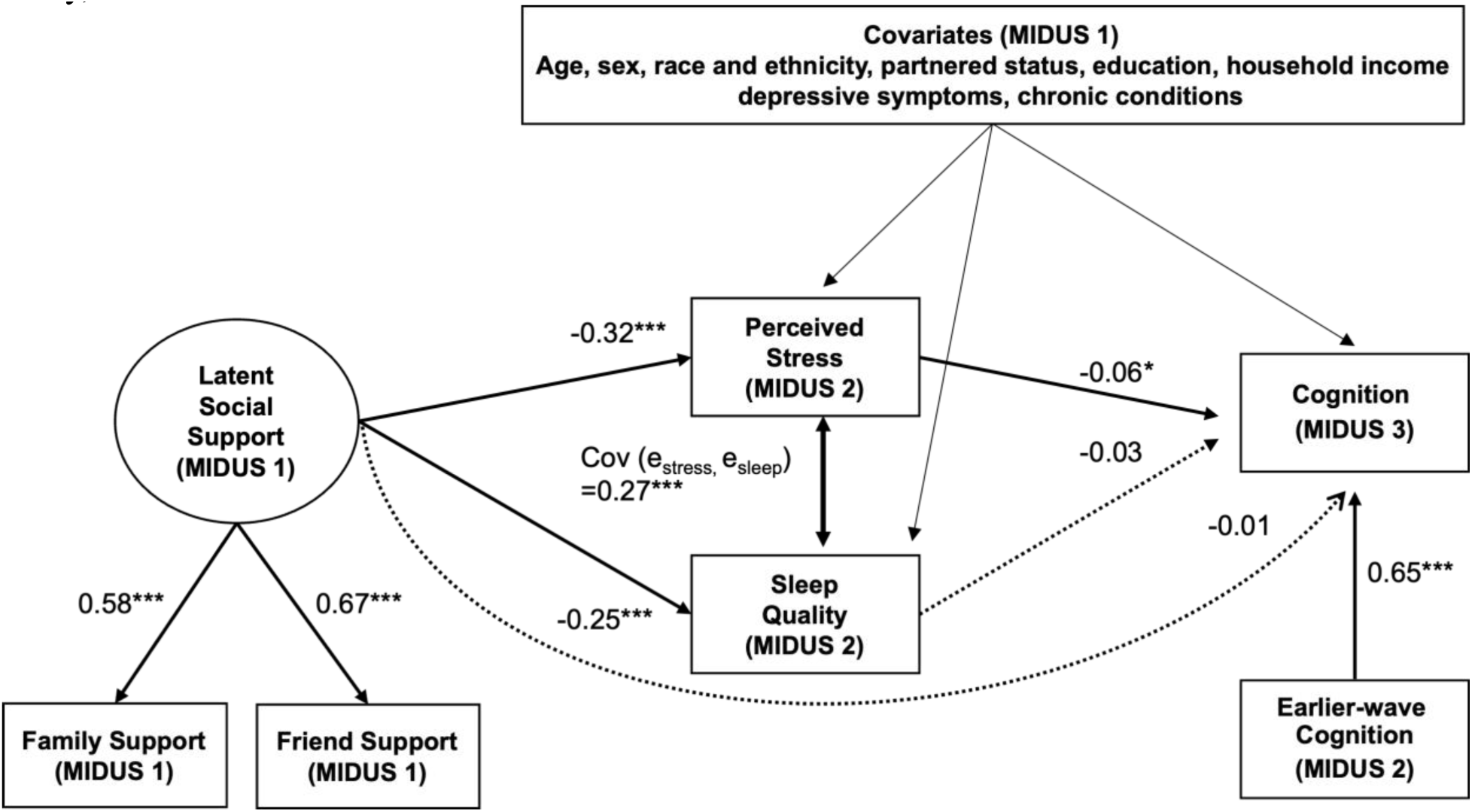
Standardized associations linking social support with cognition through perceived stress and sleep quality among adults in the United States in the Midlife in the United States study, 1995–2017. Notes: The SEM was estimated using FIML and included all 7,615 participants with data on at least one endogenous model variable. Solid lines indicate statistically significant pathways; dashed lines indicate non-significant pathways. The residual errors of perceived stress and sleep quality were allowed to covary because both mediators were assessed concurrently. Fit statistics: AIC=175,696.5, BIC=176,431.9, CD=0.90; R² for follow-up cognition=0.71. Abbreviation. MIDUS=Midlife in the United States, FIML=full information maximum likelihood, AIC=Akaike Information Criterion, BIC=Bayesian Information Criterion, CD=Coefficient of Determination.

Table 2 presents the decomposition of the direct, indirect, and total effects of social support on cognition. In the primary FIML model, the indirect effect through perceived stress was statistically significant (β=0.02, 95% CI: 0.00, 0.04), whereas the indirect effect through sleep quality was not (β=0.01, 95% CI: −0.00, 0.02). The total indirect effect was statistically significant (β=0.03, 95% CI: 0.01, 0.05), while the direct and total effects were not. The complete-case estimate for the stress-specific indirect pathway was similar in magnitude, although it was no longer statistically significant (β=0.02, 95% CI: −0.00, 0.04).

**Table 2.** Standardized direct, indirect, and total associations of social support with follow-up cognition through perceived stress and sleep quality among adults in the United States in the Midlife in the United States study, 1995–2017. Notes: Estimates are standardized product-of-coefficients effects from the primary FIML model and the complete-case sensitivity model. The primary structural equation model was estimated using maximum likelihood with missing values (full information maximum likelihood [FIML]) and robust standard errors. Standard errors and 95% confidence intervals were calculated using the delta method. The model adjusted for partnered status, age, sex, race and ethnicity, education, household income, depressive symptoms, chronic conditions, and earlier-wave cognition. n=7,615 Abbreviation. FIML=full information maximum likelihood, SE=standard error, CI=confidence interval

| Model | Effect | Standardized $\beta$ | SE | 95% CI |
| --- | --- | --- | --- | --- |
| Primary FIML | Indirect via perceived stress | 0.02 | 0.01 | 0.00, 0.04 |
|  | Indirect via sleep quality | 0.01 | 0.01 | 0.00, 0.02 |
|  | Total indirect effect | 0.03 | 0.01 | 0.01, 0.04 |
|  | Direct effect | -0.01 | 0.02 | -0.05, 0.03 |
|  | Total effect | 0.01 | 0.02 | -0.02, 0.05 |
| Complete-case | Indirect via perceived stress | 0.02 | 0.01 | 0.00, 0.04 |
|  | Indirect via sleep quality | 0.01 | 0.01 | -0.01, 0.02 |
|  | Total indirect effect | 0.03 | 0.01 | 0.00, 0.05 |
|  | Direct effect | -0.04 | 0.04 | -0.11, 0.04 |
|  | Total effect | -0.01 | 0.04 | -0.08, 0.06 |

The complete-case model re-estimated the primary analysis among participants with complete data on all modeled variables (n=738; Figure 3). In this restricted sample, the association between social support and perceived stress was slightly larger in magnitude (β=−0.33, 95% CI: −0.44, −0.22), and the path from perceived stress to cognition was attenuated but remained statistically significant (β=−0.06, 95% CI: −0.11, −0.00). The direct path from social support to cognition was not statistically significant. Model fit indices were AIC=26,201.2; BIC=26,440.6; CD=0.87; R² for follow-up cognition=0.64, and SRMR=0.02. Results were consistent when stress and sleep were tested separately as mediators in two models (Supplementary Figure 1). The perceived stress-only model showed a small significant indirect pathway, whereas the sleep quality-only indirect pathway was not statistically significant. As the sleep-only FIML model failed to converge, we estimated separate complete-case mediator-specific models.

**Figure 3.**
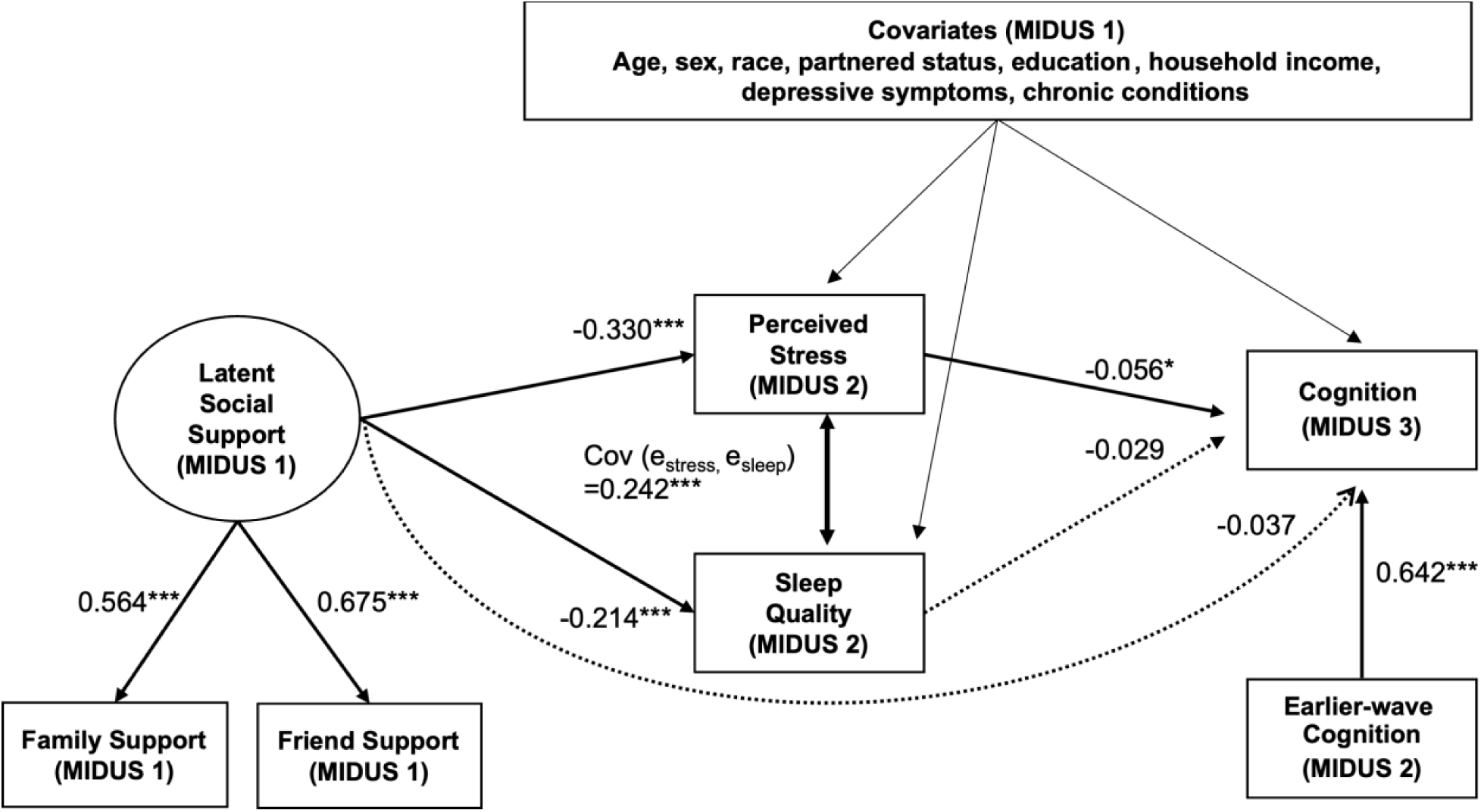
Complete-case structural equation model of social support, perceived stress, sleep quality, and cognition among adults in the United States, 1995–2017 (n=738). Notes: Values are standardized path coefficients from the complete-case sensitivity analysis restricted to participants with complete data on all modeled variables. Solid lines indicate statistical significance, while dashed lines indicate non-significant pathways. AIC = 26,201.2; BIC = 26,440.6, CD = 0.871; R² for follow-up cognition = 0.636, SRMR = 0.019 Abbreviation. AIC = Akaike Information Criterion, BIC = Bayesian Information Criterion, SRMR = Standardized Root Mean Squared Residual

Supplementary Tables 3 and 4 present the full SEM estimates for the primary FIML and complete-case models, respectively. Effect sizes were attenuated in the complete-case sample, and the stress-specific indirect effect was less precise, although the direct association between social support and cognition remained nonsignificant.

Sensitivity analyses using observed social-support measures showed a similar pattern (Supplementary Table 5). The observed family/friend composite was indirectly associated with cognition through perceived stress, whereas the sleep pathway was not significant. With family and friend support entered separately, total indirect effects were positive for both, and the stress pathway was more evident for friend support than for family support.

We estimated two sequential mediation models with alternative orderings of the concurrently assessed mediators, perceived stress and sleep quality (Supplementary Figure 2). The sequential indirect pathway was non-significant in the stress-to-sleep model (Panel A; estimate=0.01, 95% CI: −0.00, 0.02) and statistically significant in the sleep-to-stress model (Panel B; estimate=0.01, 95% CI: 0.00, 0.02). Because the mediators were measured concurrently, these estimates represent alternative statistical orderings rather than temporal sequences. The direct association between social support and cognition remained non-significant in both models.

## 4. DISCUSSION

Using approximately two decades of longitudinal MIDUS data, we found that higher perceived social support was associated with lower perceived stress and better sleep quality, but only perceived stress predicted lower subsequent cognition after adjustment for earlier-wave cognition. The model identified a small positive indirect association through perceived stress despite the absence of statistically significant total or direct associations between social support and later cognition. These findings align with the Stress Process Model and stress-buffering hypothesis, which propose that supportive relationships reduce stress responses and downstream biological dysregulation linked to cognitive aging.^10,12^ Chronic activation of stress-response systems contributes to cumulative neuroendocrine and inflammatory dysregulation, conceptualized as allostatic load, which has been implicated in cognitive aging and neurodegeneration.^11,13,14^ Together, these perspectives position perceived stress as a possible psychosocial pathway linking social support to later cognition.

Perceived stress has rarely been examined as a mediator linking social support and cognitive outcomes; prior studies have focused more on depression or loneliness.^27,28^ Unlike these constructs, perceived stress reflects appraisal of unpredictability and overload, which are central components of stress process theory.^29,30^ Our findings suggest that perceived stress was prospectively associated with later cognition independent of earlier-wave cognition and sociodemographic factors, consistent with a possible psychosocial pathway linking social support to cognitive aging.^30,31^ Although the association between perceived stress and later cognition was statistically significant, the standardized effect size was modest, suggesting that perceived stress likely represents one of multiple psychosocial and behavioral pathways contributing to cognitive aging.

Prior cross-sectional studies have identified sleep as a mediator linking social resources and cognition.^19,20^ In contrast, our longitudinal model, which adjusts for earlier-wave cognition, did not identify an independent association between sleep quality and later cognition after accounting for perceived stress. Differences in study design, population characteristics, cultural context, and measurement of sleep may contribute to the differing findings. Alternatively, sleep quality and perceived stress may be mutually related. Because both were measured concurrently at MIDUS 2, the sequential models reflect alternative statistical orderings rather than temporal sequence; they were consistent with one possible statistical parameterization in which sleep and stress were associated, but did not establish the direction of their relationship.

This study has several limitations. First, the main model was estimated using FIML under a missing-at-random assumption. However, differential attrition across MIDUS waves and selective participation in the biomarker or cognitive projects may have introduced selection bias and limited representativeness, particularly because sampling weights could not be applied across combined projects and waves. Although complete-case analyses produced similar findings, residual bias from unmeasured or missing-not-at-random mechanisms remains possible. Models did not converge among adults aged ≥65 years because of limited information, limiting the generalizability of our findings to this subgroup.

Second, models were also limited by the constructs assessed. The latent social support construct included only two indicators, resulting in a just-identified measurement model that precluded evaluation of measurement fit. Perceived stress and sleep quality were self-reported and may not reflect biological stress responses and sleep physiology, potentially introducing recall bias.^32^ Additionally, the sleep quality measure combined multiple sleep dimensions, which may have obscured dimension-specific associations. Moreover, psychosocial measures were assessed at different time points, and perceived stress and sleep quality reflected recent rather than long-term experiences. Residual confounding by other health-related and behavioral factors may remain despite adjustment for baseline chronic conditions and depressive symptoms.

Third, temporal ordering was also limited. Month-level assessment dates indicated that earlier-wave cognition preceded biomarker collection among participants with observed key variables; however, exact assessment days were unavailable and not all FIML participants had complete timing data. Temporal ordering should therefore be interpreted cautiously. Furthermore, causal inference is limited by the observational design. Although social support was associated with stress and cognition over time, these data cannot establish that social support causes changes in stress or cognition.

Nonetheless, this study has important strengths. Its longitudinal design linked social support, stress, sleep quality, and cognition across approximately two decades while accounting for earlier-wave cognition. The use of FIML and sensitivity analyses supported assessment of the robustness of the findings, although causal inference remains limited.

## 5. CONCLUSIONS

The model identified a small positive indirect association through perceived stress despite nonsignificant total and direct social support-cognition associations. Although prior research consistently shows that greater social integration and support are associated with better cognition and lower dementia risk,^33,34^ fewer longitudinal studies have examined underlying mechanisms. By modeling perceived stress and sleep quality as parallel mediators, this study extends prior work and is consistent with stress appraisal as a possible pathway linking social support to later cognitive differences. More studies are needed to determine whether strengthening supportive relationships or reducing perceived stress contributes to cognitive health.

## Declarations

### Ethics approval and consent to participate

This study used de-identified, publicly available secondary data from the Midlife in the United States (MIDUS) study. Ethical approval and informed consent were obtained by the original MIDUS investigators. No additional ethical approval or consent was required for this secondary analysis.

## Data Availability statement

Data were obtained from the publicly available Midlife in the United States study through ICPSR. Access to the underlying data is subject to the MIDUS/ICPSR data-use conditions. The analytic dataset used in this study is available from the corresponding author upon reasonable request, subject to those conditions.

## Authors’ contributions

S.R.: Conceptualization, Formal analysis, Methodology, Writing – original draft.

E.L.T.: Conceptualization, Writing – review & editing

M.B.M.: Writing – review & editing

J.A.K.: Writing – review & editing

I.S.H.: Methodology, Writing – review & editing

## Data Availability

All data produced are available online at https://midus.wisc.edu/data/index.php

---

**Supplementary Figure 1.**
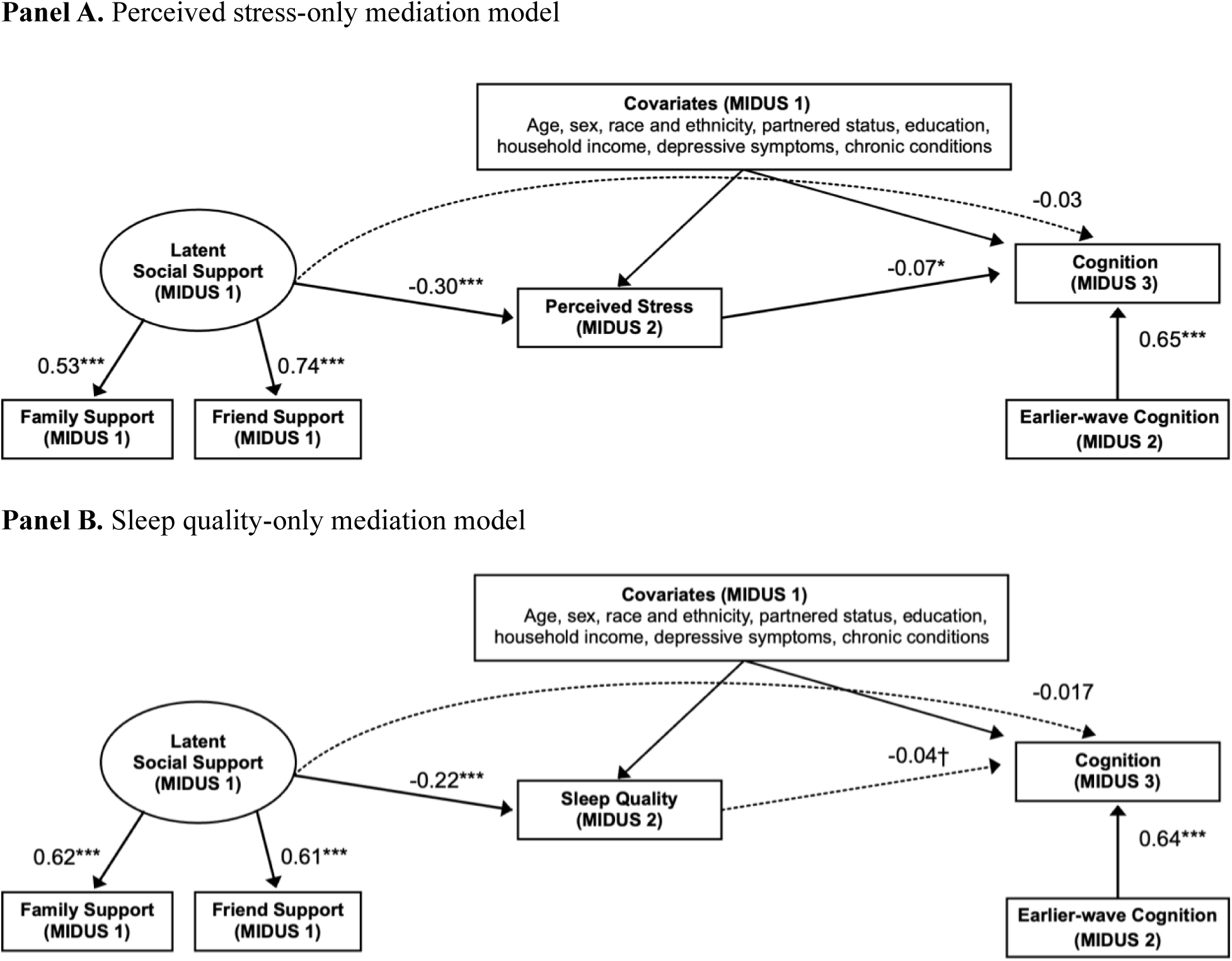
Separate complete-case mediation models of perceived stress and sleep quality among adults in the United States, 1995–2017. Notes: Values are standardized path coefficients from complete-case sensitivity analyses. Analyses for each panel were restricted to participants with complete data on all variables included in that specific model, resulting in different sample sizes across panels (Panel A: n = 773; Panel B: n = 740). Solid lines indicate statistical significance, while dashed lines indicate non-significant pathways. Panel A: AIC = 23,548.39; BIC = 23,729.75; CD = 0.876; R² for follow-up cognition = 0.638; SRMR = 0.020 Panel B: AIC = 21,662.01; BIC = 21,841.67; CD = 0.854; R² for follow-up cognition = 0.634; SRMR = 0.018 Abbreviation. MIDUS=Midlife in the United States, AIC = Akaike Information Criterion, BIC = Bayesian Information Criterion, CD = Coefficient of Determination, SRMR = Standardized Root Mean Squared Residual

**Supplementary Figure 2.**
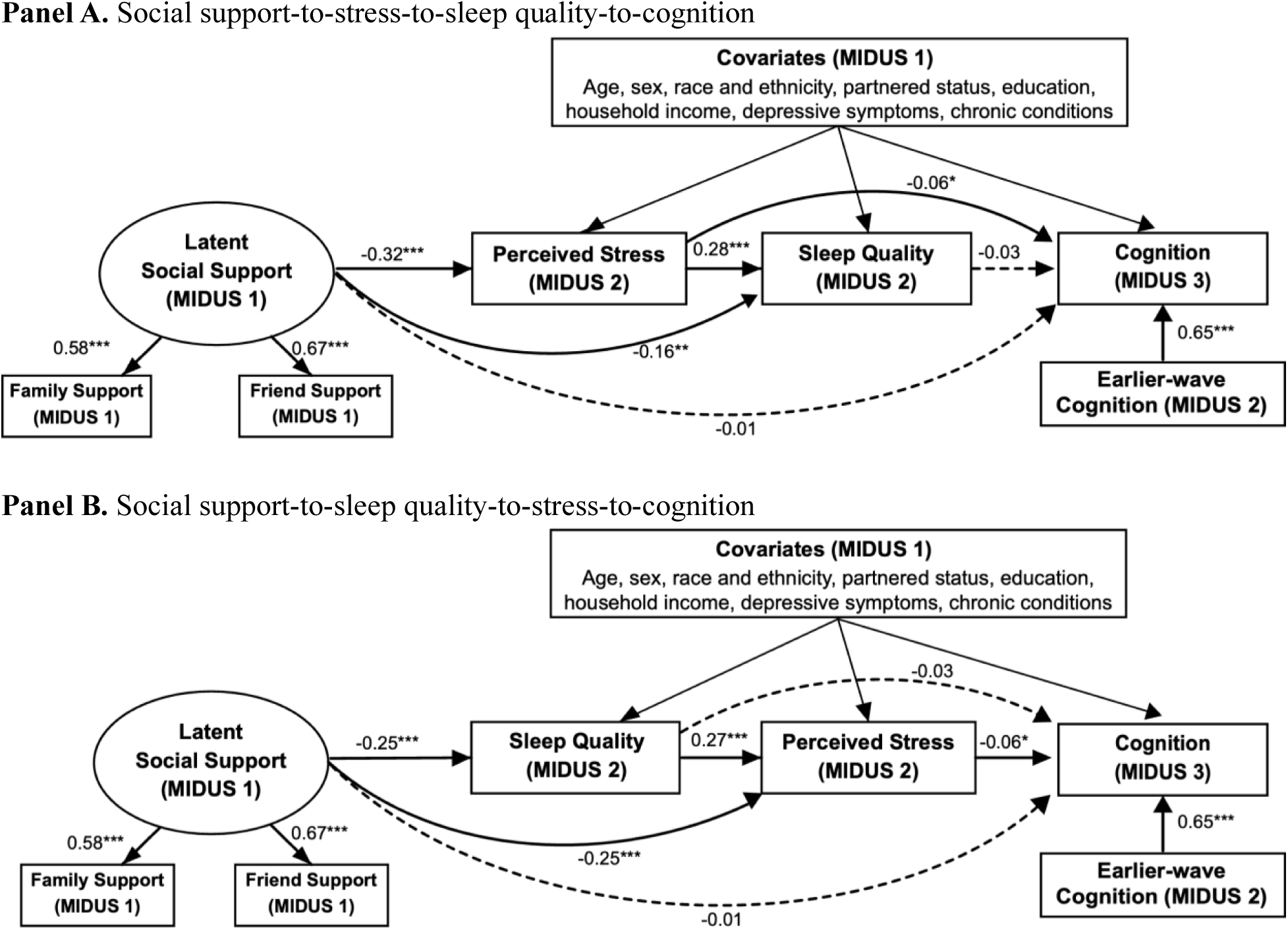
Alternative sequential mediation models among adults United States, Midlife in the United States study, 1995–2017. Notes: Values are standardized path coefficients from the primary structural equation model estimated using maximum likelihood with missing values (full information maximum likelihood [FIML]) and robust standard errors. Solid lines indicate statistical significance, while dashed lines indicate non-significant pathways. n=7,615 Fit statistics: Panel A and B: AIC=175,696.5; BIC=176,431.9; CD=0.90; R² for follow-up cognition=0.71. AIC and BIC are comparative information criteria. Abbreviation. MIDUS=Midlife in the United States, AIC=Akaike Information Criterion, BIC=Bayesian Information Criterion, CD=Coefficient of Determination.

**Supplementary Table 1.** Pairwise observed overlap among key variables for adults in the United States, Midlife in the United States study, 1995–2017. Notes: Diagonal (bold)=marginal observed N for each variable; upper triangle=N with both variables jointly observed. Social support is a latent factor indicated by family and friend support. Pairwise counts for social support reflect participants with at least one observed support indicator and the column variable observed; 6,255 had at least one support indicator, 6,233 had both indicators, and each individual support indicator had 6,244 observed values. All five key variables were jointly observed for n=743 participants (key variables only); n=738 had complete data for the complete-case sensitivity sample after adding covariates (age, sex, race and ethnicity, education, income, partnered status, depression, chronic conditions). The main FIML analytic pool comprised n=7,615 participants with data on ≥1 modeled variable.

|  | Social support | Perceived stress | Sleep quality | Earlier-wave cognition | Follow-up cognition |
| --- | --- | --- | --- | --- | --- |
| Social support | 6,255 | 1,020 | 971 | 3,806 | 2,639 |
| Perceived stress | - | 1,249 | 1,168 | 981 | 849 |
| Sleep quality | - | - | 1,172 | 935 | 809 |
| Earlier-wave cognition | - | - | - | 3,973 | 2,445 |
| Follow-up cognition | - | - | - | - | 3,043 |

**Supplementary Table 2.**
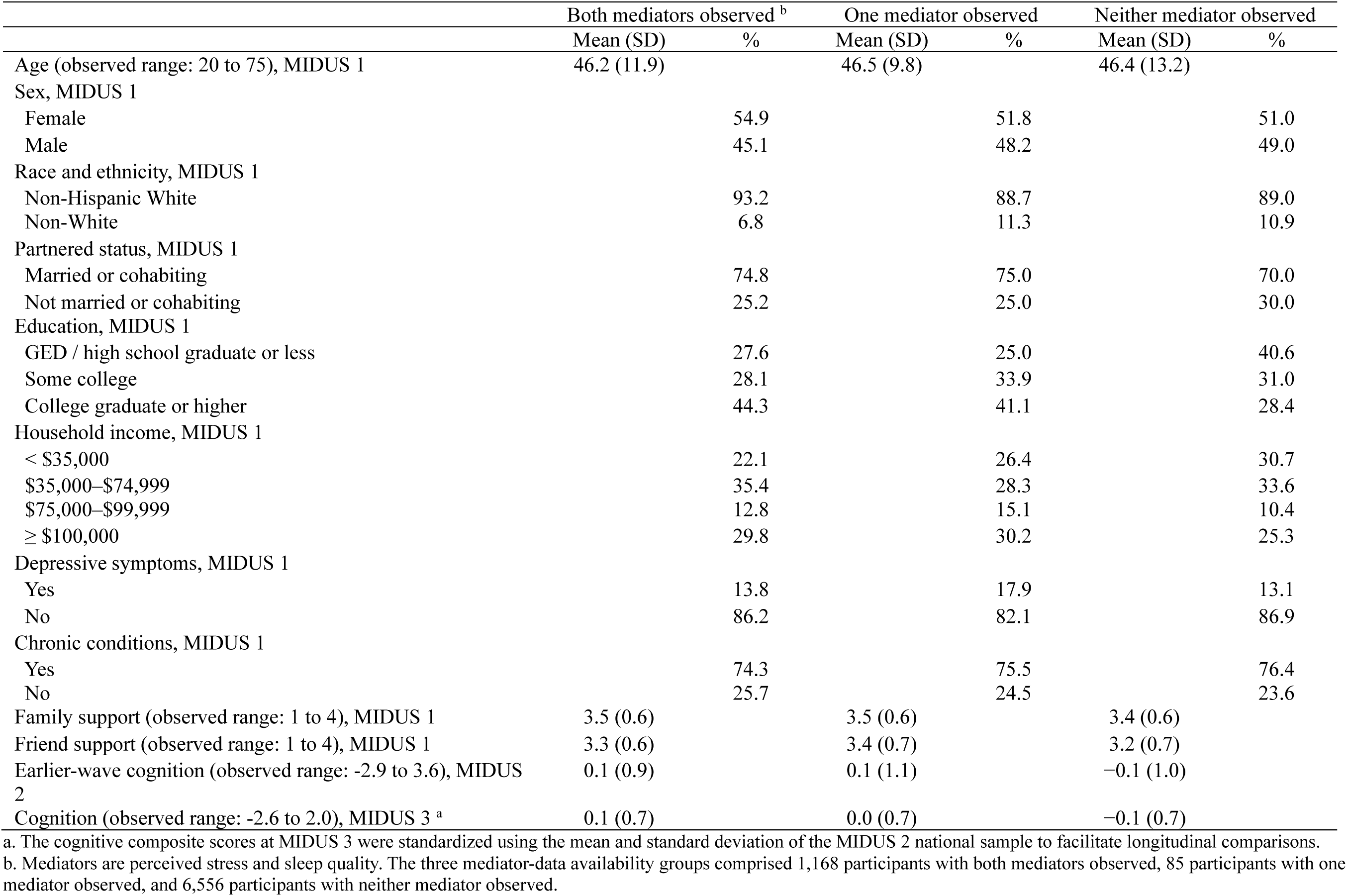
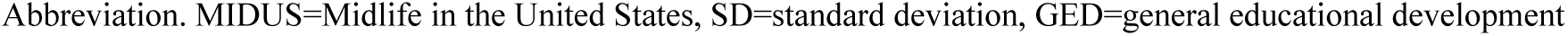
Characteristics of adults in the United States by mediator-data availability, Midlife in the United States study, 1995–2017. a. The cognitive composite scores at MIDUS 3 were standardized using the mean and standard deviation of the MIDUS 2 national sample to facilitate longitudinal comparisons. b. Mediators are perceived stress and sleep quality. The three mediator-data availability groups comprised 1,168 participants with both mediators observed, 85 participants with one mediator observed, and 6,556 participants with neither mediator observed. Abbreviation. MIDUS=Midlife in the United States, SD=standard deviation, GED=general educational development

|  | Both mediators observed <sup>b</sup> |  | One mediator observed |  | Neither mediator observed |  |
| --- | --- | --- | --- | --- | --- | --- |
|  | Mean (SD) | % | Mean (SD) | % | Mean (SD) | % |
| Age (observed range: 20 to 75), MIDUS 1 | 46.2 (11.9) |  | 46.5 (9.8) |  | 46.4 (13.2) |  |
| Sex, MIDUS 1 |  |  |  |  |  |  |
| Female |  | 54.9 |  | 51.8 |  | 51.0 |
| Male |  | 45.1 |  | 48.2 |  | 49.0 |
| Race and ethnicity, MIDUS 1 |  |  |  |  |  |  |
| Non-Hispanic White |  | 93.2 |  | 88.7 |  | 89.0 |
| Non-White |  | 6.8 |  | 11.3 |  | 10.9 |
| Partnered status, MIDUS 1 |  |  |  |  |  |  |
| Married or cohabiting |  | 74.8 |  | 75.0 |  | 70.0 |
| Not married or cohabiting |  | 25.2 |  | 25.0 |  | 30.0 |
| Education, MIDUS 1 |  |  |  |  |  |  |
| GED / high school graduate or less |  | 27.6 |  | 25.0 |  | 40.6 |
| Some college |  | 28.1 |  | 33.9 |  | 31.0 |
| College graduate or higher |  | 44.3 |  | 41.1 |  | 28.4 |
| Household income, MIDUS 1 |  |  |  |  |  |  |
| < \$35,000 | | 22.1 | | 26.4 | | 30.7 |
| \$35,000–\$74,999 | | 35.4 | | 28.3 | | 33.6 |
| \$75,000–\$99,999 | | 12.8 | | 15.1 | | 10.4 |
| ≥ \$100,000 | | 29.8 | | 30.2 | | 25.3 |
| Depressive symptoms, MIDUS 1 |  |  |  |  |  |  |
| Yes |  | 13.8 |  | 17.9 |  | 13.1 |
| No |  | 86.2 |  | 82.1 |  | 86.9 |
| Chronic conditions, MIDUS 1 |  |  |  |  |  |  |
| Yes |  | 74.3 |  | 75.5 |  | 76.4 |
| No |  | 25.7 |  | 24.5 |  | 23.6 |
| Family support (observed range: 1 to 4), MIDUS 1 | 3.5 (0.6) |  | 3.5 (0.6) |  | 3.4 (0.6) |  |
| Friend support (observed range: 1 to 4), MIDUS 1 | 3.3 (0.6) |  | 3.4 (0.7) |  | 3.2 (0.7) |  |
| Earlier-wave cognition (observed range: -2.9 to 3.6), MIDUS 2 | 0.1 (0.9) |  | 0.1 (1.1) |  | –0.1 (1.0) |  |
| Cognition (observed range: -2.6 to 2.0), MIDUS 3 <sup>a</sup> | 0.1 (0.7) |  | 0.0 (0.7) |  | –0.1 (0.7) |  |

**Supplementary Table 3.**
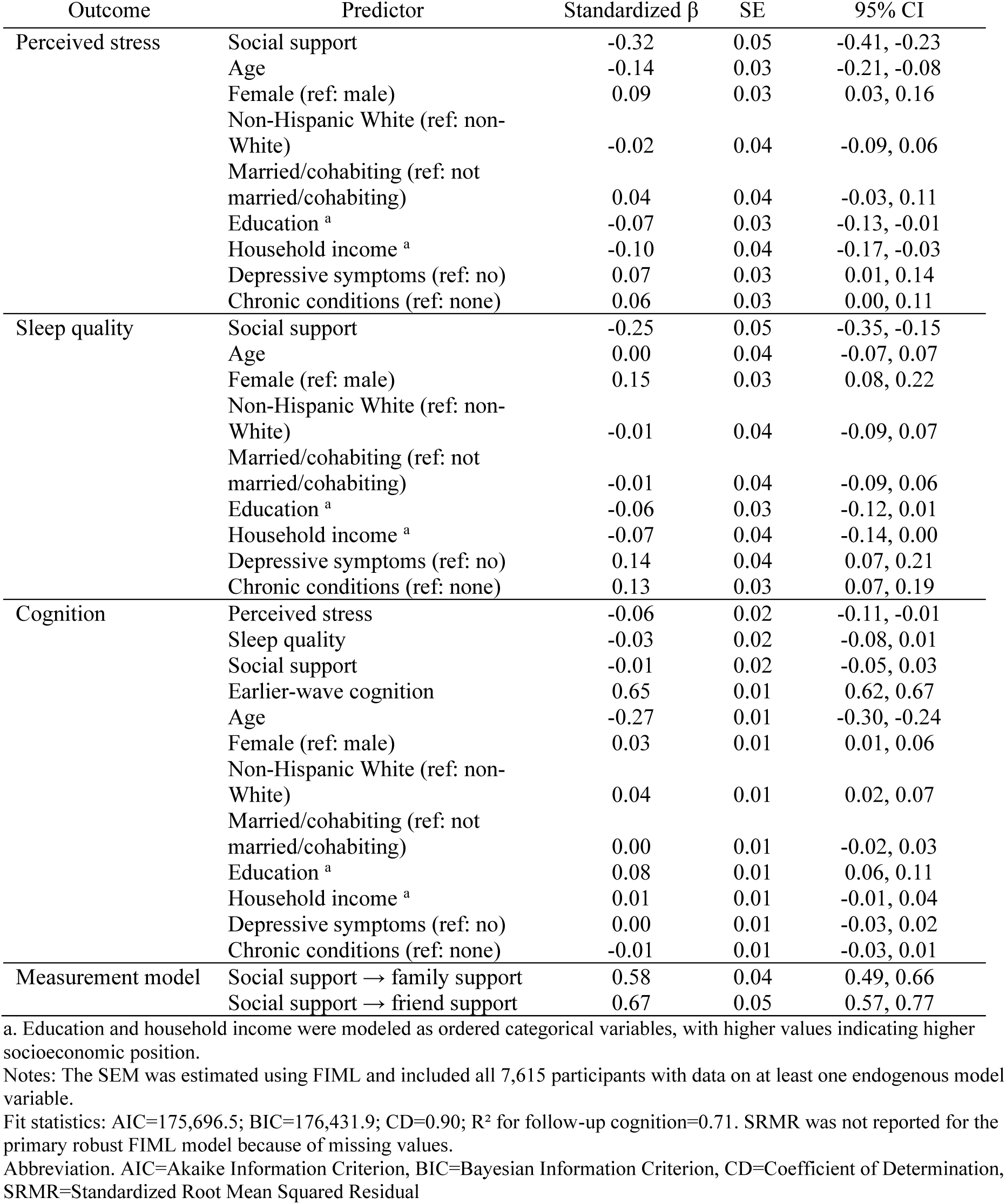
Standardized structural equation model estimates among adults in the United States, Midlife in the United States study, 1995–2017. a. Education and household income were modeled as ordered categorical variables, with higher values indicating higher socioeconomic position. Notes: The SEM was estimated using FIML and included all 7,615 participants with data on at least one endogenous model variable. Fit statistics: AIC=175,696.5; BIC=176,431.9; CD=0.90; R² for follow-up cognition=0.71. SRMR was not reported for the primary robust FIML model because of missing values. Abbreviation. AIC=Akaike Information Criterion, BIC=Bayesian Information Criterion, CD=Coefficient of Determination, SRMR=Standardized Root Mean Squared Residual

| Outcome | Predictor | Standardized $\beta$ | SE | 95% CI |
| --- | --- | --- | --- | --- |
| Perceived stress | Social support | -0.32 | 0.05 | -0.41, -0.23 |
|  | Age | -0.14 | 0.03 | -0.21, -0.08 |
|  | Female (ref: male) | 0.09 | 0.03 | 0.03, 0.16 |
|  | Non-Hispanic White (ref: non-White) | -0.02 | 0.04 | -0.09, 0.06 |
|  | Married/cohabiting (ref: not married/cohabiting) | 0.04 | 0.04 | -0.03, 0.11 |
|  | Education <sup>a</sup> | -0.07 | 0.03 | -0.13, -0.01 |
|  | Household income <sup>a</sup> | -0.10 | 0.04 | -0.17, -0.03 |
|  | Depressive symptoms (ref: no) | 0.07 | 0.03 | 0.01, 0.14 |
|  | Chronic conditions (ref: none) | 0.06 | 0.03 | 0.00, 0.11 |
| Sleep quality | Social support | -0.25 | 0.05 | -0.35, -0.15 |
|  | Age | 0.00 | 0.04 | -0.07, 0.07 |
|  | Female (ref: male) | 0.15 | 0.03 | 0.08, 0.22 |
|  | Non-Hispanic White (ref: non-White) | -0.01 | 0.04 | -0.09, 0.07 |
|  | Married/cohabiting (ref: not married/cohabiting) | -0.01 | 0.04 | -0.09, 0.06 |
|  | Education <sup>a</sup> | -0.06 | 0.03 | -0.12, 0.01 |
|  | Household income <sup>a</sup> | -0.07 | 0.04 | -0.14, 0.00 |
|  | Depressive symptoms (ref: no) | 0.14 | 0.04 | 0.07, 0.21 |
|  | Chronic conditions (ref: none) | 0.13 | 0.03 | 0.07, 0.19 |
| Cognition | Perceived stress | -0.06 | 0.02 | -0.11, -0.01 |
|  | Sleep quality | -0.03 | 0.02 | -0.08, 0.01 |
|  | Social support | -0.01 | 0.02 | -0.05, 0.03 |
|  | Earlier-wave cognition | 0.65 | 0.01 | 0.62, 0.67 |
|  | Age | -0.27 | 0.01 | -0.30, -0.24 |
|  | Female (ref: male) | 0.03 | 0.01 | 0.01, 0.06 |
|  | Non-Hispanic White (ref: non-White) | 0.04 | 0.01 | 0.02, 0.07 |
|  | Married/cohabiting (ref: not married/cohabiting) | 0.00 | 0.01 | -0.02, 0.03 |
|  | Education <sup>a</sup> | 0.08 | 0.01 | 0.06, 0.11 |
|  | Household income <sup>a</sup> | 0.01 | 0.01 | -0.01, 0.04 |
|  | Depressive symptoms (ref: no) | 0.00 | 0.01 | -0.03, 0.02 |
|  | Chronic conditions (ref: none) | -0.01 | 0.01 | -0.03, 0.01 |
| Measurement model | Social support → family support | 0.58 | 0.04 | 0.49, 0.66 |
|  | Social support → friend support | 0.67 | 0.05 | 0.57, 0.77 |

Fit statistics: AIC=175,696.5; BIC=176,431.9; CD=0.90; R<sup>2</sup> for follow-up cognition=0.71. SRMR was not reported for the primary robust FIML model because of missing values.

**Supplementary Table 4.** Complete-case standardized structural equation model estimates among adults in the United States, Midlife in the United States study, 1995–2017. a. Education and household income were modeled as ordered categorical variables, with higher values indicating higher socioeconomic position. Notes: Values are standardized path coefficients from the complete-case sensitivity analysis restricted to participants with complete data on all modeled variables (n=738). Robust standard errors and 95% confidence intervals are reported. Fit statistics: AIC=26,201.2; BIC=26,440.6; CD=0.87; SRMR=0.02; R² for follow-up cognition=0.64. Abbreviation. SE=standard error, CI=confidence interval, CD=Coefficient of Determination, SRMR=Standardized Root Mean Squared Residual

| Outcome | Predictor | Standardized $\beta$ | SE | 95% CI |
| --- | --- | --- | --- | --- |
| Perceived stress | Social support | -0.33 | 0.05 | -0.44, -0.22 |
|  | Age | -0.17 | 0.04 | -0.23, -0.10 |
|  | Female (ref: male) | 0.11 | 0.04 | 0.04, 0.19 |
|  | Non-Hispanic White (ref: non-White) | 0.02 | 0.03 | -0.04, 0.08 |
|  | Married/cohabiting (ref: not married/cohabiting) | 0.06 | 0.04 | -0.02, 0.15 |
|  | Education <sup>a</sup> | -0.02 | 0.04 | -0.10, 0.05 |
|  | Household income <sup>a</sup> | -0.11 | 0.04 | -0.19, -0.02 |
|  | Depressive symptoms (ref: no) | 0.08 | 0.04 | 0.00, 0.16 |
|  | Chronic conditions (ref: none) | 0.04 | 0.03 | -0.03, 0.10 |
| Sleep quality | Social support | -0.21 | 0.06 | -0.32, -0.11 |
|  | Age | -0.06 | 0.04 | -0.12, 0.01 |
|  | Female (ref: male) | 0.16 | 0.04 | 0.08, 0.23 |
|  | Non-Hispanic White (ref: non-White) | 0.01 | 0.03 | -0.05, 0.08 |
|  | Married/cohabiting (ref: not married/cohabiting) | 0.01 | 0.04 | -0.07, 0.09 |
|  | Education <sup>a</sup> | -0.03 | 0.04 | -0.10, 0.05 |
|  | Household income <sup>a</sup> | -0.06 | 0.04 | -0.14, 0.01 |
|  | Depressive symptoms (ref: no) | 0.15 | 0.04 | 0.07, 0.23 |
|  | Chronic conditions (ref: none) | 0.13 | 0.03 | 0.06, 0.20 |
| Cognition | Perceived stress | -0.06 | 0.03 | -0.11, 0.00 |
|  | Sleep quality | -0.03 | 0.03 | -0.08, 0.02 |
|  | Social support | -0.04 | 0.04 | -0.11, 0.04 |
|  | Earlier-wave cognition | 0.64 | 0.02 | 0.60, 0.69 |
|  | Age | -0.23 | 0.02 | -0.28, -0.18 |
|  | Female (ref: male) | 0.02 | 0.03 | -0.02, 0.07 |
|  | Non-Hispanic White (ref: non-White) | 0.04 | 0.02 | -0.01, 0.08 |
|  | Married/cohabiting (ref: not married/cohabiting) | -0.01 | 0.03 | -0.06, 0.04 |
|  | Education <sup>a</sup> | 0.10 | 0.03 | 0.05, 0.14 |
|  | Household income <sup>a</sup> | 0.02 | 0.03 | -0.03, 0.07 |
|  | Depressive symptoms (ref: no) | -0.03 | 0.02 | -0.08, 0.02 |
|  | Chronic conditions (ref: none) | -0.03 | 0.02 | -0.07, 0.02 |
| Measurement model | Social support → family support | 0.56 | 0.07 | 0.42, 0.71 |
|  | Social support → friend support | 0.68 | 0.10 | 0.49, 0.86 |

Fit statistics: AIC=26,201.2; BIC=26,440.6; CD=0.87; SRMR=0.02; R<sup>2</sup> for follow-up cognition=0.64.

**Supplementary Table 5.**
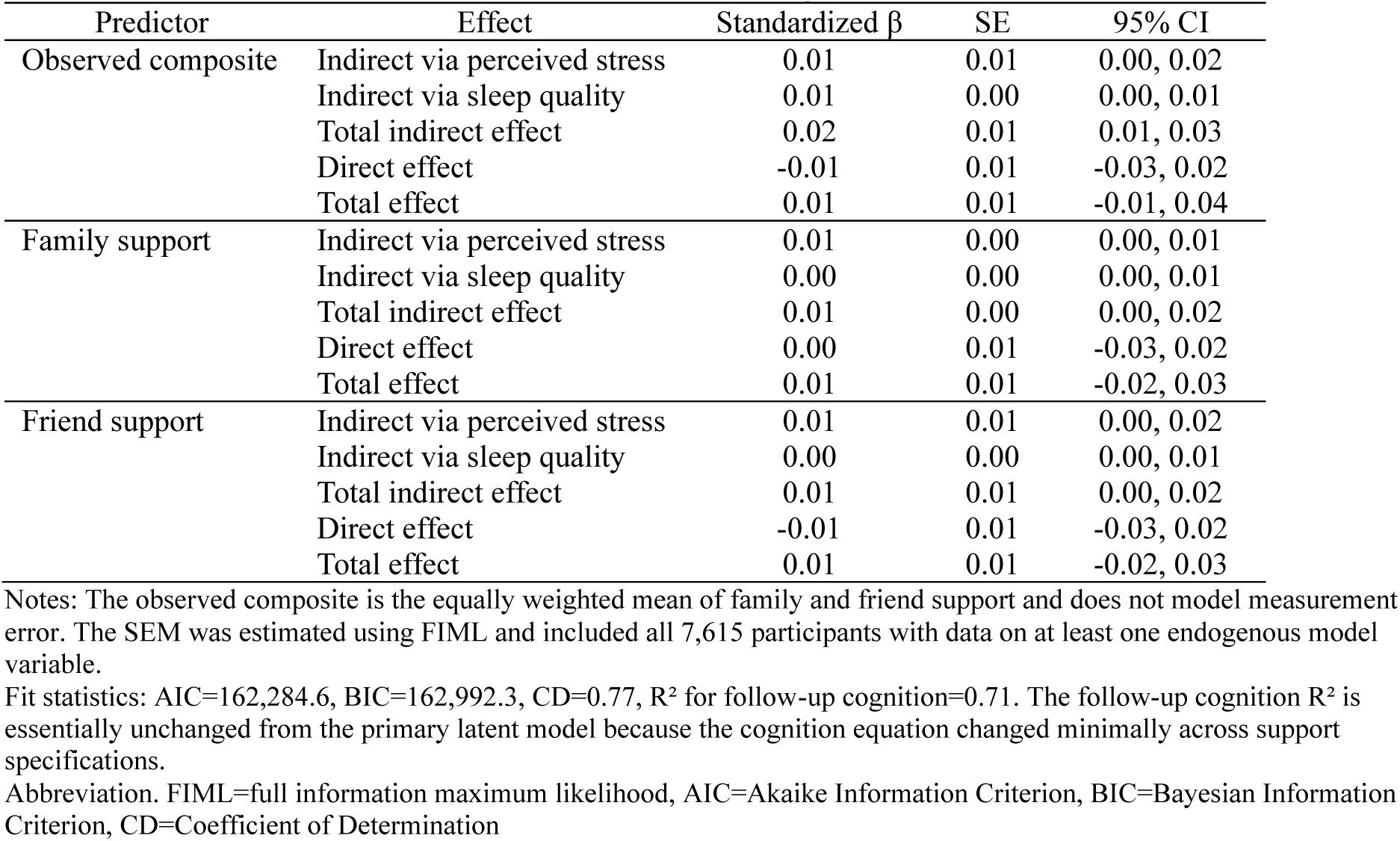
Sensitivity analysis of support specifications and cognition among adults in the United States, Midlife in the United States study, 1995–2017. Notes: The observed composite is the equally weighted mean of family and friend support and does not model measurement error. The SEM was estimated using FIML and included all 7,615 participants with data on at least one endogenous model variable. Fit statistics: AIC=162,284.6, BIC=162,992.3, CD=0.77, R² for follow-up cognition=0.71. The follow-up cognition R² is essentially unchanged from the primary latent model because the cognition equation changed minimally across support specifications. Abbreviation. FIML=full information maximum likelihood, AIC=Akaike Information Criterion, BIC=Bayesian Information Criterion, CD=Coefficient of Determination

